# High frequency, high throughput quantification of SARS-CoV-2 RNA in wastewater settled solids at eight publicly owned treatment works in Northern California shows strong association with COVID-19 incidence

**DOI:** 10.1101/2021.07.16.21260627

**Authors:** Marlene K. Wolfe, Aaron Topol, Alisha Knudson, Adrian Simpson, Bradley White, Duc J. Vugia, Alexander T. Yu, Linlin Li, Michael Balliet, Pamela Stoddard, George S. Han, Krista R. Wigginton, Alexandria B. Boehm

## Abstract

A number of recent retrospective studies have demonstrated that SARS-CoV-2 RNA concentrations in wastewater are associated with COVID-19 cases in the corresponding sewersheds. Implementing high-resolution, prospective efforts across multiple plants depends on sensitive measurements that are representative of COVID-19 cases, scalable for high throughput analysis, and comparable across laboratories. We conducted a prospective study across eight publicly owned treatment works (POTWs). A focus on SARS-CoV-2 RNA in solids enabled us to scale-up our measurements with a commercial lab partner. Samples were collected daily and results were posted to a website within 24-hours. SARS-CoV-2 RNA in daily samples correlated to incidence COVID-19 cases in the sewersheds; a 1 log_10_ increase in SARS-CoV-2 RNA in settled solids corresponds to a 0.58 log_10_ (4X) increase in sewershed incidence rate. SARS-CoV-2 RNA signals measured with the commercial laboratory partner were comparable across plants and to measurements conducted in a university laboratory when normalized by pepper mild mottle virus PMMoV RNA. Results suggest that SARS-CoV-2 RNA should be detectable in settled solids for COVID-19 incidence rates > 1/100,000 (range 0.8 - 2.3 cases per 100,000). These sensitive, representative, scalable, and comparable methods will be valuable for future efforts to scale-up wastewater-based epidemiology.

**Importance:** Access to reliable, rapid monitoring data is critical to guide response to an infectious disease outbreak. For pathogens that are shed in feces or urine, monitoring wastewater can provide a cost-effective snapshot of transmission in an entire community via a single sample. In order for a method to be useful for ongoing COVID-19 monitoring, it should be sensitive for detection of low concentrations of SARS-CoV-2, representative of incidence rates in the community, scalable to generate data quickly, and comparable across laboratories. This paper presents a method utilizing wastewater solids to meet these goals, producing measurements of SARS-CoV-2 RNA strongly associated with COVID-19 cases in the sewershed of a publicly owned treatment work. Results, provided within 24 hrs, can be used to detect incidence rates as low as approximately 1/100,000 cases and can be normalized for comparison across locations generating data using different methods.

## Introduction

The COVID-19 pandemic has prompted the rapid and widespread maturation of wastewater based epidemiology (WBE). Through the first year of the pandemic, retrospective analyses of wastewater samples for SARS-CoV-2 RNA demonstrated strong correlations between SARS-CoV-2 target concentrations and infection incidence ^1–8^. Wastewater monitoring can therefore provide a cost-effective snapshot of transmission in a community by using just one sample to provide information that is unbiased by access to testing or symptom status. To be of most value for public health officials, the methods used to produce data should be 1) sensitive to detect low levels of viral RNA in wastewater, 2) representative of the disease rates in the community, 3) scalable to provide high-throughput results with a short turnaround time, and 4) comparable between labs and different approaches.

Numerous independent COVID-19 WBE efforts have covered monitoring at a range of scales, from the building level to country-wide implementation ^7,9,10^. The methods employed have also varied, with the vast majority focusing on the liquid fraction of municipal wastewater. Due to the dilute nature of SARS-CoV-2 RNA in wastewater, most methods that focus on influent require a number of preanalytical steps to concentrate SARS-CoV-2 prior to extracting the viral RNA. Depending on the method, these steps include ultrafiltration ^2,5^, organic flocculation coupled with centrifugation ^3,11^, and charged membrane filtration ^12^. Each of these listed concentration steps add significant time, equipment, and personnel requirements to SARS-CoV-2 RNA quantification in wastewater. For effective monitoring, methods need to be scalable. SARS-CoV-2 RNA measurements should be prospective as opposed to retrospective, should provide high-resolution (e.g. daily) data on SARS-CoV-2 levels, and should result in data within hours or days of sample collection. As wastewater monitoring expands to more communities, methods focused on quantifying SARS-CoV-2 in liquid are not readily amenable to scale-up and automation due to the additional steps needed to prepare the samples.

The majority of SARS-CoV-2 RNA in wastewater originates in the feces of infected individuals. Even after mixing with wastewater liquid, coronaviruses have a stronger affinity to the solid fraction of wastewater, even higher than the affinities of nonenveloped viruses ^13^. Studies comparing SARS-CoV-2 RNA concentrations in the liquid and solid fractions of real wastewater have demonstrated that the solids harbor 3-4 orders of magnitude higher concentrations of SARS-CoV-2 RNA on a per mass basis than liquid influent ^3,14^. As a result, the settling of wastewater solids that happens during primary treatment at most wastewater treatment plants serves as a built-in concentration step for wastewater monitoring. SARS-CoV-2 RNA quantification from these samples simultaneously requires less preanalytical processing and, on a per mass basis, will result in higher measured concentrations than measurements made on the liquid fraction of wastewater. If plants do not have a primary clarifier or samples are taken at the sub-sewershed level, solids may still be concentrated from influent using standard methods ^15^. SARS-CoV-2 monitoring focused on settled solids may therefore be both more sensitive and more conducive to scale-up and automation.

In this study, we demonstrate that a SARS-CoV-2 monitoring project focused on wastewater solids can be readily scaled to produce sensitive results that are representative of COVID-19 incidence through a high-frequency effort conducted through a commercial laboratory with <24 hour turn-around times. We initiated a prospective monitoring project across eight POTWs in the greater San Francisco Bay and Sacramento areas of California to measure SARS-CoV-2 RNA in daily samples. Results were consistently reported to stakeholders and agencies within 24 hours of sample collection. Here we show that SARS-CoV-2 RNA in daily samples correlates strongly to COVID-19 incidence rates in the sewersheds. Results indicate SARS-CoV-2 RNA signals are comparable across plants and across laboratories using different measurement methods. We present the empirical sensitivity of the measurements to incidence rates in the sewersheds.

## Methods

A wastewater monitoring program was initiated in November 2020 to implement methods previously developed for analysis of wastewater solids ^3^ in a high-throughput format for high-resolution (daily) data collection at 8 publicly owned treatment works (POTWs) in the greater San Francisco Bay and Sacramento areas of California. Whereas the original protocol took several days to complete, the high throughput protocol uses liquid handling robots and other automations to process samples in less than 24 h. Protocols used for sample processing as described below are publicly available on protocols.io ^16–18^, and results of wastewater measurements have been provided to stakeholders in real time on a website visualizing the data (wbe.stanford.edu). Four POTWs serve populations of Santa Clara County, CA (SJ, Gil, Sun, and PA), and the others each serve a portion of San Mateo County (SVCW, PA), San Francisco County (Ocean), Sacramento County (Sac), and Yolo County (Dav). The eight POTWs serve between 66,000 and 1,500,000 residents and have permitted flows between 8.5 and 181 million gallons per day (Table 1).

**Table 1.** Description of POTWs and samples in this study. Start date is in 2020. N in the last column is the number of samples analyzed at each POTW. Information on the residence time of sewage in the network and solids in the clarifier was obtained through a survey completed by POTW operators and managers.

| Acronym | County | Served population | Permitted flow (MGD) | Pre-treatment? | Solids collected from | Residence time of solids in clarifier (h) | Residence time in the sewer network (h) | Grab / composite? | Start date (N) |
| --- | --- | --- | --- | --- | --- | --- | --- | --- | --- |
| SJ | Santa Clara | 1,458,017 | 167 | FeCl<sub>3</sub> | Primary clarifier | 1-2 | 4-18 | C | 15 Nov (137) |
| PA | Santa Clara | 213,968 | 39 |  | Primary clarifier | 2-3 | 2-9 | G | 16 Nov (135) |
| Sun | Santa Clara | 169,000 | 29.5 |  | Primary clarifier | 3-6 | 6-12 | G | 30 Nov (122) |
| Gil | Santa Clara | 110,338 | 8.5 |  | Influent | NA | 3-6 | C | 30 Nov (121) |
| SVCW | San Mateo | 220,000 | 29 | FeCl<sub>3</sub> | Primary clarifier | 2-3 | 2-8 | G | 8 Dec (114) |
| Ocean | San Francisco | 250,000 | 43 |  | Primary clarifier | 3-6 | 6-12 | G | 8 Dec (104) |
| Dav | Yolo | 66,622 | 7.5 |  | Primary clarifier | 2 | 2 | G | 23 Nov (127) |
| Sac | Sacramento | 1,480,000 | 181 |  | Primary clarifier | 1 | 15 | G | 8 Dec (114) |

### Sample Collection

Samples were collected by POTW staff using sterile technique in clean, labeled bottles provided by our team. POTWs were not provided additional compensation for participation. Approximately 50 ml of settled solids were collected each day from each sewage treatment plant between mid-to-late November 2020 and March 31, 2021. At 7 of the 8 POTWs, settled solids were collected from the primary clarifier; at these POTW the residence time of solids in the primary clarifier ranged from 1 to 6 h (Table 1). Settled solids samples were grab samples at all plants except for SJ. At SJ, POTW staff manually collected a 24 h composite sample ^3^ (Table 1). At Gil, solids were settled from a 24 h composite influent sample using standard method 160.5 ^15^. Samples were immediately stored at 4°C and transported to the lab where processing began immediately (within 6 hours of collection).

### Sample Preparation

The solids were dewatered by centrifugation at 24,000 x g for 30 minutes at 4°C. The supernatant was aspirated and discarded. A 0.5 - 1 g aliquot of the dewatered solids was dried at 110°C for 19-24 hrs to determine its dry weight. Bovine coronavirus (BCoV) was used as a positive recovery control. Each day, attenuated bovine coronavirus vaccine (PBS Animal Health, Calf-Guard Cattle Vaccine) was spiked into DNA/RNA shield solution (Zymo Research) at a concentration of 1.5 µL /mL. Dewatered solids were resuspended in the BCoV-spiked DNA/RNA shield to a concentration of 75 mg/mL. This concentration of solids was chosen as previous work titrated solutions with varying concentrations of solids to identify a concentration at which inhibition of the SARS-CoV-2 assays was minimized (data not shown). Between five and ten 5/32” Stainless Steel Grinding Balls (OPS Diagnostics) were added to each sample which was subsequently homogenized by shaking with a Geno/Grinder 2010 (Spex SamplePrep). Samples were then briefly centrifuged to remove air bubbles introduced during the homogenization process, and then vortexed to re-mix the sample.

### RNA Extraction

RNA was extracted from 10 replicate aliquots per sample. For each replicate, RNA was extracted from 300 µl of homogenized sample using the Chemagic™ Viral DNA/RNA 300 Kit H96 for the Perkin Elmer Chemagic 360 followed by PCR Inhibitor Removal with the Zymo OneStep-96 PCR Inhibitor Removal Kit. Extraction negative controls (water) and extraction positive controls were extracted using the same protocol as the homogenized samples. The positive controls consisted of 500 copies of SARS-CoV-2 genomic RNA (ATCC® VR-1986D™) in the BCoV-spiked DNA/RNA shield solution described above.

### Droplet Digital PCR

RNA extracts were used as template in digital droplet RT-PCR assays for SARS-CoV-2 N, S, and ORF1a RNA gene targets in a triplex assay, and BCoV and PMMoV in a duplex assay (see Table S1 for primer and probe sequences, purchased from IDT). PMMoV is highly abundant in human stool and domestic wastewater globally ^19,20^ and is used here as an internal recovery and fecal strength control. Undiluted extract was used for the SARS-CoV-2 assay template and a 1:100 dilution of the extract was used for the BCoV / PMMoV assay template. Digital RT-PCR was performed on 20 µl samples from a 22 µl reaction volume, prepared using 5.5 µl template, mixed with 5.5 µl of One-Step RT-ddPCR Advanced Kit for Probes (Bio-Rad 1863021), 2.2 µl Reverse Transcriptase, 1.1 µl DTT and primers and probes at a final concentration of 900 nM and 250 nM respectively. Droplets were generated using the AutoDG Automated Droplet Generator (Bio-Rad). PCR was performed using Mastercycler Pro with the following protocol: reverse transcription at 50°C for 60 minutes, enzyme activation at 95°C for 5 minutes, 40 cycles of denaturation at 95°C for 30 seconds and annealing and extension at either 59°C (for SARS-CoV-2 assay) or 56°C (for PMMoV/BCoV duplex assay) for 30 seconds, enzyme deactivation at 98°C for 10 minutes then an indefinite hold at 4°C. The ramp rate for temperature changes were set to 2°C/second and the final hold at 4°C was performed for a minimum of 30 minutes to allow the droplets to stabilize. Droplets were analyzed using the QX200 Droplet Reader (Bio-Rad). All liquid transfers were performed using the Agilent Bravo (Agilent Technologies).

Each sample was run in 10 replicate wells, extraction negative controls were run in 7 wells, and extraction positive controls in 1 well. In addition, PCR positive controls for SARS-CoV-2 RNA, BCoV, and PMMoV were run in 1 well, and NTC were run in 7 wells. Positive controls consisted of BCoV and PMMoV gene block controls (dsDNA purchased from IDT) and gRNA of SARS-CoV-2 (ATCC® VR-1986D™). Results from replicate wells were merged for analysis. Thresholding was done using QuantaSoft™ Analysis Pro Software (Bio-Rad, version 1.0.596). Additional details are provided in supporting material per the dMIQE guidelines ^21^. In order for a sample to be recorded as positive, it had to have at least 3 positive droplets. Three positive droplets corresponds to a concentration between ∼500-1000 cp/g; the range in values is a result of the range in the equivalent mass of dry solids added to the wells.

Concentrations of RNA targets were converted to concentrations per dry weight of solids in units of copies/g dry weight using dimensional analysis. The total error is reported as standard deviations and includes the errors associated with the poisson distribution and the variability among the 10 replicates. The recovery of BCoV was determined by normalizing the concentration of BCoV by the expected concentration given the value measured in the spiked DNA/RNA shield. BCoV was used solely as a process control; samples were rerun in cases where the recovery of BCoV was less than 1%.

### Ancillary wastewater data

Wastestream influent total suspended solids (TSS) concentrations in mg/L data were obtained from POTW staff. If TSS measurements were not coincident on the day that a sample was taken, the TSS value for that day was estimated using linear interpolation.

### COVID-19 case data

Counts of laboratory-confirmed COVID-19 incident cases as a function of episode date (earliest of reported symptom onset, laboratory result, or case record create dates) for each sewershed were obtained from local or state sources through data-use agreements. Case data were aggregated within the sewersheds based on georeferenced reported home addresses, which were delineated using the POTW-specific GIS shape files. COVID-19 incidence rates per 100,000 population were calculated using the estimated population served in each sewershed.

### Statistical analysis

Statistics were computed using RStudio (version 1.1.1073). For ddRT-PCR data, results are reported both daily and using a 7-day, centered trimmed moving average where the largest and smallest values are dropped prior to averaging. COVID-19 incidence data are represented in terms of 7-day, centered running average (smoothed) new cases for each of the sewersheds. The latter is justified owing to the “weekend effect” associated with a reduction in test seeking behavior, testing availability, and result reporting ^22^. Hereafter, any reference to incidence or incidence rate will refer to the value from the 7-day smoothed average.

Incidence rates were compared to SARS-CoV-2 RNA concentrations, SARS-CoV-2 RNA concentrations normalized by PMMoV concentrations (C_PMMoV_), and SARS-CoV-2 RNA concentrations scaled by the factor F= K_dp_(1+K_d_TSS)/(C_PMMoV_K_d_(1+K_dp_TSS)) where K_d_ and K_dp_ are the partitioning coefficients for SARS-CoV-2 and PMMoV, respectively, and the other terms have been defined. The scaling factor falls from a mass balance model relating the number of SARS-CoV-2 fecal shedders in a sewershed to the concentration of SARS-CoV-2 RNA in settled solids ^7^ and will hereafter be referred to as F for simplicity. In applying F to the data, we used K_d_ = 1000 and K_dp_ = 100 ^7^. Results were similar when the analysis was repeated with varying K_d_ and K_dp_ between 100 and 10^4^ ml/g (data not shown).

Non-parametric Kendall’s tau and Kruskal-Wallis methods were used to test hypotheses regarding associations and trends as data were neither normally nor log-normally distributed based on Shapiro-Wilk tests. Linear regressions were used to assess slopes describing relationships between incidence rates and N gene concentrations normalized by PMMoV for each POTW and for all POTWs aggregated. To account for variability of wastewater measurements, Kendall’s tau empirical p-values and regression coefficients m were determined using 1,000 bootstrap re-samplings that incorporate non-detect measurements and measurement errors ^3^, and median tau, regression coefficients m, standard error, R^2^, and empirical p are reported. For trimmed data, bootstrapping utilized 95% confidence intervals accounting for variability in the 5 measurements included in the trimmed average.

The minimum incidence rate at which wastewater solids contain measurable SARS-CoV-2 RNA was estimated for each POTW. Predictions were calculated within the bootstrapping approach by using coefficients from the observed linear relationship between log_10_-transformed COVID-19 incidence rate and log_10_ N cp/g measured in wastewater solids for each bootstrapped sample to predict the incidence rate when wastewater solids contained 750 cp/g of the N gene. The median predicted incidence rate and interquartile range reported. The 750 cp/g value was chosen because it falls in the middle of the range of the lowest detectable concentration of the method.

Concentrations of SARS-CoV-2 N1 and PMMoV targets in settled solids reported by Wolfe et al. ^7^ collected early in the pandemic (Spring-Fall 2020) at seven POTWs including two from this project (SJ and Ocean) were acquired along with associated sewershed COVID-19 incidence rates. These data were used to assess whether trends observed with data in the present study are distinct from those reported by Wolfe et al. ^7^. The empirical regression coefficients for the two data sets were compared using Kruskal Wallis test to test the null hypothesis of no difference between the distribution of coefficients. Those authors measured N1 and PMMoV using a different workflow than used in the present study. The detection limit of N1 reported by those authors was ∼40 cp/g; in assessing associations using these data, the bootstrapping approach samples from a uniform distribution defined by 0 and 40 cp/g to assign a value to non-detects in the data set.

## Results

### QA/QC

Negative and positive extraction and PCR controls were negative and positive, respectively. BCoV recoveries were, on average 57% (standard deviation = 39%) and all were above 1% (Figure S1). Sample standard deviations for the SARS-CoV-2, PMMoV, and BCoV recovery quantification estimated from the merged wells were, on average 19%, 19%, and 14% of the measurement. As the samples were extracted ten times and each extract analyzed in one of 10 replicate wells which were merged, the replicate variability incorporates variation from both RNA extraction and RT-ddPCR with a heterogeneous solids sample. Additional reporting according to dMIQE guidelines are in the SI. Wastewater data used in this study are available through the Stanford Digital Repository ^23^.

### POTW adherence to protocols

Adherence to daily sampling was high among POTWs. During the duration of this study, POTW staff were unable to collect samples on 0 days (4 POTW), 1 day (2 POTW), 2 days (1 POTW), and 10 days (1 POTW). The reason for missed samples was most often lack of sampling bottles or miscommunication with new staff. For the POTW that missed 10 samples, samples were missed mostly on Sundays and holidays when the POTW had limited staff on site.

### Measurement overview

Across all samples, PMMoV ranged from 4.3 × 10^7^ to 7.1 × 10^9^ cp/g (average = 6.6 × 10^8^). PMMoV was different between POTW (Kruskal-Wallis p<10^−15^) with Ocean tending to have the lowest PMMoV and Gil the highest (Figure S2). SARS-CoV-2 RNA gene concentrations ranged from 630 to 3.7×10^6^ (N gene), ND to 3.2×10^6^ (S gene) and ND to 3.0×10^6^ (ORF1a) cp/g across all samples. A total of 4 samples returned non-detects during this period (1 for S and 3 for ORF1a) and the value of 300 cp/g (approx half the detection limit) was substituted for these for further analysis. N, S, and ORF1a gene concentrations were highly correlated (r_p_ ranged from 0.97 to 0.98 for log_10_-transformed data aggregated across plants). Pairwise linear regressions between log_10_-transformed N, S, and ORF1a concentrations returned slopes ∼1 (Figure S3). Results with untransformed variables were similar (see SI). Therefore, further analyses in this paper focuses on the N gene alone.

### Relationship between SARS-CoV-2 RNA in solids and incident cases

Concentrations of SARS-CoV-2 RNA rose over the “winter surge” in COVID-19 incidence in November and December of 2020 and declined to lower levels at the end of March (Figure 1). There is an apparent dip in incidence rate at the peak of the surge; this was probably due to decreased test seeking behavior and reduced testing availability during the week between Christmas and New Year’s Day. During the time period of this study there were no days on which any POTW achieved a non-detect across the three genes, nor were there days where incident case numbers in the sewershed were 0. Over the duration of the study, the lowest smoothed incident rates observed across the eight POTWs ranged from 1.9 to 8.5 cases per 100,000 people (at Dav and Sac, respectively). These correspond to the low numbers of daily smoothed incident cases ranging from 1.3 to 125.7 in these two sewersheds. Wastewater data availability preceded case data availability; case reporting delays in this region at the time were greater than 3 days.

**Fig 1.**
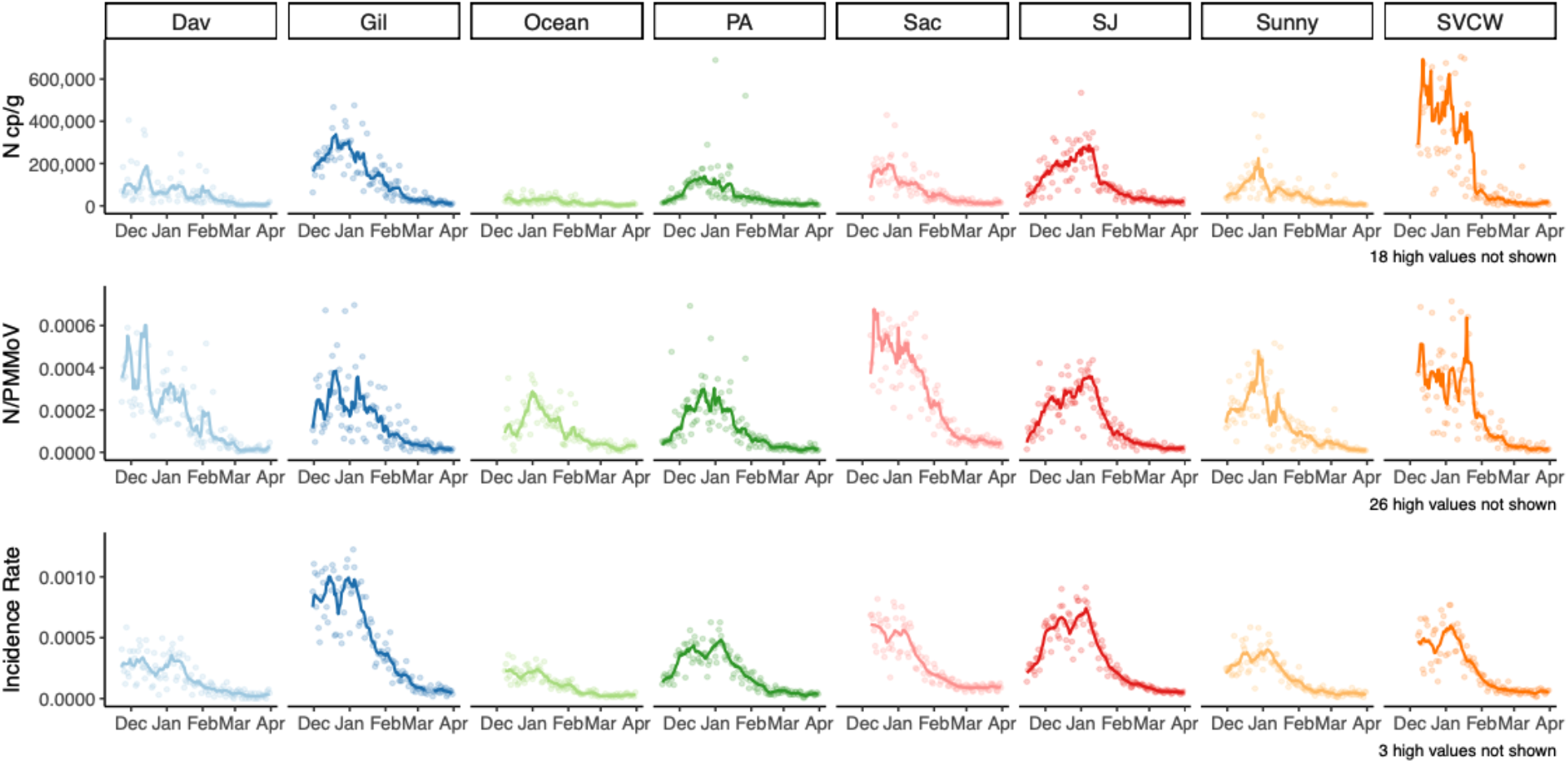
Time series of (top to bottom) N cp/g in wastewater, N/PMMoV, and laboratory-confirmed SARS-CoV-2 incidence rate (bottom) for each of 8 POTWs and their sewersheds from mid-November/December 2020 - March 31 2021. Points represent daily values. Lines are 7-day, centered smoothed averages. For wastewater, this is a trimmed average where the highest and lowest values from the seven day period are removed before averaging.

Kendall’s tau between 7-d smoothed incidence rates and N gene concentrations ranged from 0.46 (Ocean) to 0.70 (SJ) (all p<0.001) (Table 2, Figure S4-S6). Within POTW, associations were significantly enhanced when N was normalized by PMMoV, or scaled by F at 5 of the 8 POTW, although the effect size was small (effect size < 0.1, Kruskal Wallis p<0.001); at Gil, Sun, and SVCW, the association was significantly weakened (effect size < 0.1, Kruskal Wallis p<0.001). When 7-d trimmed average N gene concentrations were used in lieu of raw N gene concentrations the significance and direction of differences in association between N, N normalized by PMMoV and N scaled by F and incidence rate were unchanged at each plant. Kendall’s tau was significantly higher for each of these measurements when data was trimmed, although the effect size was small (effect size <0.1, Kruskal Wallis all p<10^−15^).

**Table 2.** Empirical Kendall’s tau for the test of the null hypothesis that the wastewater variable is not associated with the 7-d smoothed new COVID-19 incidence rate in the sewershed the day of the wastewater measurement. Values are shown for each sewershed for both daily values and 7-day trimmed wastewater values for N cp/g, N normalized by PMMoV. Empirical p<0.001 for all.

| Kendall's Tau |  |  |  |  |  |  |
| --- | --- | --- | --- | --- | --- | --- |
|  | N cp/g |  | N/PMMoV |  | F*N |  |
| Plant | Daily | Trimmed | Daily | Trimmed | Daily | Trimmed |
| All | 0.657 | 0.684 | 0.585 | 0.601 | 0.586 | 0.601 |
| SJ | 0.632 | 0.650 | 0.648 | 0.680 | 0.648 | 0.674 |
| PA | 0.699 | 0.743 | 0.709 | 0.739 | 0.718 | 0.740 |
| Sun | 0.565 | 0.618 | 0.560 | 0.618 | 0.557 | 0.612 |
| Gil | 0.697 | 0.721 | 0.575 | 0.646 | 0.578 | 0.648 |
| SVCW | 0.643 | 0.660 | 0.608 | 0.553 | 0.605 | 0.553 |
| Ocean | 0.456 | 0.477 | 0.546 | 0.510 | 0.530 | 0.499 |
| Dav | 0.637 | 0.662 | 0.640 | 0.664 | 0.641 | 0.665 |
| Sac | 0.595 | 0.605 | 0.641 | 0.657 | 0.640 | 0.656 |

When data from the eight POTWs are aggregated, a strong association between wastewater concentrations of SARS-CoV-2 RNA and incidence rate persists (Figure 2). Kendall’s tau between raw N concentrations and incidence rates aggregated across plants is 0.66 (p<0.001); tau decreases to 0.58 (p<10^−15^) when data are normalized by PMMoV or scaled by F (Table 2, Figure S7). Associations between incidence rate and 7-d trimmed average wastewater data are similar to those between incidence rate and raw wastewater data (Table 2). Linear regressions between COVID-19 incidence rate and wastewater values suggest that for a 1 log_10_ increase in N cp/g, there is a 0.59 (± 0.01 standard error) log_10_ increase in incidence rate (R^2^ = 0.67), and for N normalized by PMMoV and N scaled by F there is a 0.58 (±0.02) log_10_ increase in COVID-19 incidence (R^2^ = 0.58).

**Fig 2.**
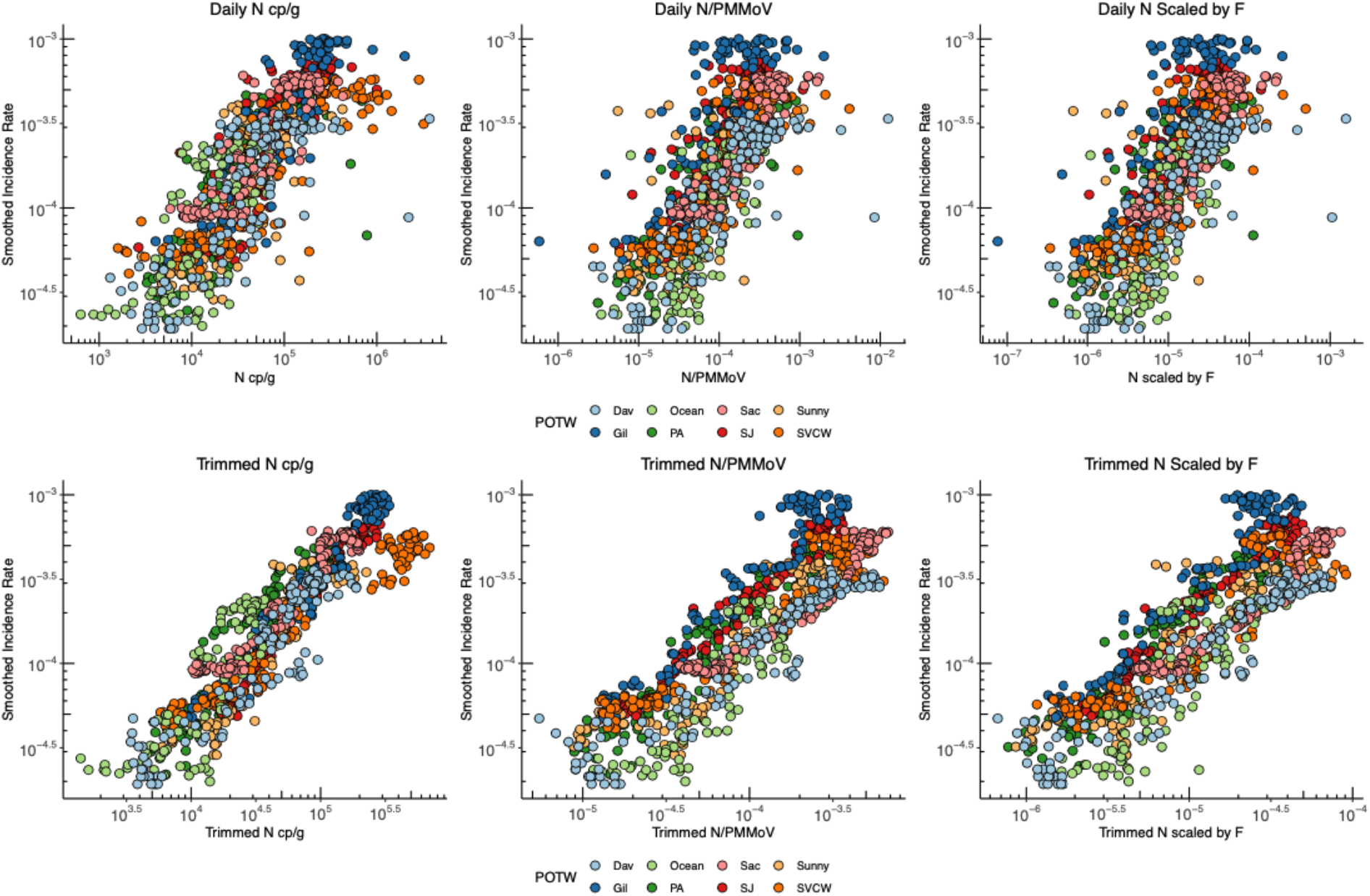
7-day smoothed COVID-19 incidence rate plotted against daily wastewater measurements (top row) and 7-day trimmed wastewater measurements (bottom row). From left to right, plots show the association between incidence rate and 1) N gene copies/g, 2) N gc/g normalized by PMMoV, 3) N gc/g scaled by a factor (F) that includes PMMoV, TSS and partitioning coefficients presented in Wolfe et al. ^7^.

Incidence rates and concentrations of the N gene in settled solids reported by Wolfe et al. ^7^ were added to the aggregated plots (Figure 3). Those data were generated in a different laboratory using a different method from those used for the data reported herein, and represent data from diverse POTWs in California, Illinois, and New York during a different phase of the pandemic (Spring-Fall 2020). When displayed as raw N gene concentrations versus incidence rate, the Wolfe et al. data do not fall on the same data cloud as the data generated in the present study and the slope of the linear regression lines through each data set are divergent (slope = 0.13 ± 0.03 and 0.59 ± 0.01 log_10_ incidence rate/log_10_ N cp/g for Wolfe et al. and the data from this paper, respectively). However, when all the data are normalized by PMMoV or scaled by a factor that includes TSS and partitioning coefficients, the Wolfe et al. data collapse onto the data generated in this study. After normalizing by PMMoV or scaling by F, the regression lines for the two methods remain significantly different (Kruskal Wallis p<0.001 for all), however the slopes describing the relationship between wastewater values and incidence rate are similar between the two methods after applying these approaches (slope = 0.24 ± 0.03 and 0.58 ± 0.02 log_10_ incidence rate/log_10_ wastewater value for both normalized and scaled data from Wolfe et al. and this paper, respectively).

**Fig 3.**
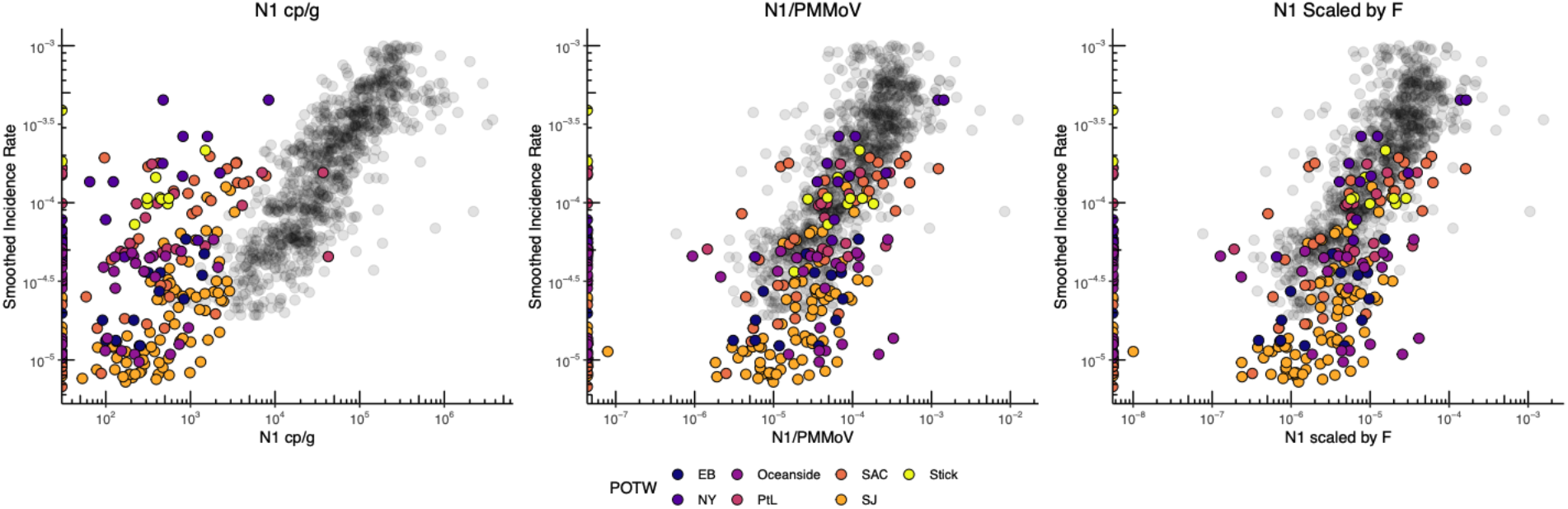
Data from Wolfe et al 2021 (in color) showing daily measurement of SARS-CoV-2 in wastewater overlaid on results from Fig x showing daily measurements from this study. Both datasets show 7-day smoothed COVID-19 incidence rate plotted against daily wastewater measurements 1) N gene copies/g, 2) N gc/g normalized by PMMoV, 3) N gc/g scaled by F. The two datasets are both generated from analysis of solids, however Wolfe et al used a more labor-intensive, small scale set of methods and slightly different genomic targets vs the high-throughput methods presented in this paper.

### Empirical detection limits

The linear relationship between log_10_ -transformed incidence rate and log_10_ -transformed N cp/g for each plant was used to estimate the incidence rate detection limit assuming an assay detection limit of 750 cp/g. The minimum number of estimated cases detectable in each sewershed was calculated based on the population served by each POTW. Across the eight POTWs, the average estimated incidence rate detection limit was 1.4 cases per 100,000 people (range 0.8 - 2.3 cases per 100,000 depending on POTW; Table 3). In the POTW service areas included in this project, these rates correspond to between 1.0 and 26.3 cases per sewershed depending on sewershed (Dav and Sac are represented by the minimum and maximum report in the range, respectively). This estimation is corroborated by measurement of N gene at concentrations above 750 cp/g when there were 1.3 recorded cases at Dav although data in this study does not reach the detection limit and future data can lend more insight to these predictions.

**Table 3.** Coefficients from bootstrapped regression models describing the relationship between log_10_-transformed COVID-19 incidence and log_10_-transformed N cp/g for each plant and the lowest estimated incidence rate and number of cases expected to result in positive wastewater result. Estimated incidence rate and incidence are shown as the median predicted value from bootstrapping with the 25th and 75th percentiles in parentheses. The observed range of 7-day smoothed incidence rate and number of cases during the study period is also shown. Intcpt is the intercept.

|  | Intcpt | Slope | R<sup>2</sup> | Population | Lowest<br>Estimated<br>Incidence Rate<br>(cases/100,000) | Lowest<br>Estimated<br>Number<br>Cases | Range of<br>Observed<br>Incidence Rate<br>(cases/100,000) | Range of<br>Observed<br>Number of<br>Cases |
| --- | --- | --- | --- | --- | --- | --- | --- | --- |
| SJ | -7.21 | 0.74 | 0.75 | 1,458,017 | 0.8 (0.8-0.8) | 11.8 (11.5-12.2) | 4.9-74.2 | 71.1-1081.7 |
| PA | -6.66 | 0.63 | 0.62 | 213,968 | 1.4 (1.3-1.4) | 2.9 (2.9-3.0) | 2.2-48.3 | 4.7-103.4 |
| Sun | -6.73 | 0.62 | 0.61 | 169,000 | 1.1 (1.1-1.2) | 1.9 (1.9-2.0) | 2.9-40.5 | 4.9-68.4 |
| Gil | -7.12 | 0.74 | 0.80 | 110,338 | 1.0 (1.0-1.0) | 1.1 (1.0-1.1) | 4.8-100.2 | 5.3-110.6 |
| SVCW | -5.91 | 0.44 | 0.75 | 220,000 | 2.3 (2.3-2.3) | 5.0 (5.0-5.1) | 4.1-60.1 | 9.0-132.1 |
| Ocean | -6.46 | 0.58 | 0.46 | 250,000 | 1.6 (1.5-1.7) | 4.0 (3.9-4.2) | 2.0-24.6 | 5.0-61.4 |
| Dav | -6.31 | 0.52 | 0.76 | 66,622 | 1.6 (1.5-1.6) | 1.0 (1.0-1.1) | 1.9-35.8 | 1.3-23.9 |
| Sac | -6.50 | 0.61 | 0.61 | 1,480,000 | 1.8 (1.7-1.8) | 26.3 (25.6-27.0) | 8.6-61.1 | 127.4-904.0 |

## Discussion

Outbreak monitoring through clinical data alone can present challenges - for diseases where individuals may be infectious before onset of symptoms or asymptomatic such as COVID-19 and norovirus, among others, tests may not be sought or may be unavailable. During the COVID-19 pandemic, racial and ethnic minority groups in the United States have been at higher risk of morbidity and mortality in part due to lack of access to testing and care^24^. Wastewater monitoring for SARS-CoV-2 RNA can provide an estimate of COVID-19 incidence rates in communities that are unbiased by these factors. For example, apparent dips in incidence rates associated with testing bias during the holidays were not reflected in wastewater trends. We modified an academic laboratory protocol for measuring SARS-CoV-2 RNA in wastewater solids so that it could be executed quickly (in less than 24 hours) and with numerous samples in parallel. Using the high-throughput and rapid protocol, we provided daily measurements of SARS-CoV-2 RNA and associated quality assurance control metrics to stakeholders on a daily basis via a public website (wbe.stanford.edu). Over the period of more than 4 months represented in this analysis, sample results were always available within 24 h of sample retrieval from the POTW with the exception of holidays. POTW staff rarely missed sample collection.

The strong association between SARS-CoV-2 RNA in solids and incident case rates within sewersheds, and across sewersheds is striking. The nearly linear relationships between these measures, whether they are examined for individual POTW or data aggregated across all POTW, provide evidence that measurements of SARS-CoV-2 RNA in wastewater solids reflect trends in COVID-19 incidence rates for the sewershed. Importantly, associations are strong for all POTWs and there is no difference between plants that provided grab samples of settled solids, composite samples of settled solids, or solids settled from composite influent samples. This suggests that even for plants without a primary clarifier, solids represent a viable matrix for wastewater surveillance efforts. While 750 cp/g is the estimated detection limit for the methods used for this data, the detection limit can be lowered even further with some adjustments to these methods.

As evidenced by the strong relationship between POTW-aggregated SARS-CoV-2 RNA gene concentrations and incidence rate, adjusting for POTW-specific characteristics (like fecal strength, flow rate, TSS, for example) is not necessary to arrive at an empirical relation between the two variables for the POTWs included in this study. While removing outliers from the wastewater data using a trimmed average approach improves the visual association between wastewater measurements and incidence rates, the statistical measures of association (Kendall’s tau) do not differ substantially between trimmed averages and are used in lieu of raw gene concentrations. Similarly, normalizing by PMMoV or scaling the SARS-CoV-2 RNA concentrations by a factor accounting for PMMoV, TSS, and virus partitioning (F) does not improve the strength of the associations. Because TSS is so similar across POTWs (Table S2) and the same partitioning coefficients were used in calculation F for each POTW, it is not surprising that the scaling N gene concentrations by F does not appreciably affect associations with incidence rate; this was also reported by Wolfe et al. ^7^.

We suggest that SARS-CoV-2 RNA gene concentrations should be normalized by PMMoV and a trimmed average applied for stakeholder and public consumption of the data. Normalizing by PMMoV serves a number of purposes: (1) it adjusts for variable viral RNA recovery between samples (assuming PMMoV RNA recovery is similar to that of SARS-CoV-2 RNA), (2) it adjusts for differences in fecal strength of the wastestream, and (3) the normalization falls from a mass balance model relating the number of shedders to concentrations in wastewater solids ^7^. Although normalizing by PMMoV did not improve the associations observed in data from this study, the benefit of this approach is especially realized in our analysis illustrating that results from two distinct laboratories using distinct methods to measure SARS-CoV-2 RNA in solids can be combined when SARS-CoV-2 RNA concentrations are normalized by PMMoV. In this case, the median recovery of BCoV spiked into the solids described by Wolfe et al. ^7^ was 4% while BCoV recovery in the present study was approximately 10 times higher. As such, normalizing by PMMoV likely served to adjust for variable recovery. Additionally, recent work by Simpson et al. ^25^ suggests that normalizing by PMMoV can serve to correct for degradation of SARS-CoV-2 RNA during sample storage.

A benefit of having daily measurements is the ability to apply a trimmed averaging approach for data visualization. We recommend applying a trimmed average to eliminate the influence of outliers during data visualization by public health professionals and non-experts, including the public. Environmental data exhibit variability that is caused by different factors than those that are most familiar to these audiences, e.g. biases that cause variability in clinical case or syndromic data. SARS-CoV-2 RNA concentrations in wastewater may exhibit high-frequency variability for several reasons. Among the most influential are: 1) changing contributions to the sample from sudden movement of people shedding viral RNA in their stool into or out of the sewershed or intermittent deliveries of septic waste that could vary in content or time represented relative to wastewater flows, 2) variability in fecal shedding rates from person to person ^26,27^, and 3) samples are spatially heterogenous and, although our samples are well mixed, inhomogeneities could exist at spatial scales greater than those captured by our sampling.

## Conclusions

We measured three SARS-CoV-2 RNA targets in wastewater solids daily for over 4 months for consumption by POTW and public health stakeholders. The methods used were shown to be 1) sensitive to identify low concentrations of SARS-CoV-2 and COVID-19 incidence rates in the associated sewersheds, 2) scalable to a high-throughput format with results delivered daily within 24 hrs, 3) representative of disease incidence in the sewersheds served, and 4) comparable across laboratories using different methods to analyze solids. POTW staff provided samples and rarely missed a sample. Using a high-throughput method that takes advantage of automation and robotics, we were able to provide sample results within 24 hours of sample receipt and displayed those results on a public website for stakeholders. As the use of wastewater monitoring is poised to scale globally for monitoring not just COVID-19, but also other diseases, it is critical that methods are scalable for use by labs producing reliable results at an industrial scale. Strict QA/QC procedures (including negative and positive extraction and PCR controls for all targets, and recovery controls) coupled to replicate analyses (n=10) for each sample ensured high quality data. Measurements at each POTW are strongly associated with lab-confirmed COVID-19 incidence rates in the sewersheds dated to the time of specimen collection. Further, the strong association persists when data are aggregated across POTW, suggesting that SARS-CoV-2 RNA concentrations in settled solids from different POTW can be directly compared to infer relative incidence rates across POTW sewersheds. Although normalizing data by PMMoV was not necessary for comparing measurements made at different POTW in this study, we show how normalizing by PMMoV can allow for data from wastewater solids measured by different methods and labs to be readily compared to each other. Public health representatives have expressed an interest in comparing these data across sewersheds. Future work will utilize a longer time series of daily wastewater measurements from these POTWs to investigate the appropriate cadence of sampling to capture incident case trends during different phases of the pandemic, and utilize solids settled from samples capturing sub-sewershed areas to illustrate use of these methods at different scales.

## Data Availability

Wastewater data used for this analysis are available through the Stanford Digital Repository.

https://purl.stanford.edu/bx987vn9177

## Acknowledgments

This work is supported by a gift from the CDC-Foundation. The graphical abstract was created with BioRender.com. Numerous people contributed to sample collection and case data acquisition including Srividhya Ramamoorthy (Sac), Michael Cook (Sac), Ursula Bigler (Sac), James Noss (Sac), Lisa C. Thompson (Sac), Payak Sarkar (SJ), Noel Enoki (SJ), and Amy Wong (SJ), Alexandre Miot (Ocean), Lily Chan (Ocean), the Oceanside plant operations personnel, Karin North (PA), Armando Guizar (PA), Saeid Vaziry (Gil), Chris Vasquez (Gil), Alo Kauravlla (Sun), Maria Gawat (SVCW), Tiffany Ishaya (SVCW), Eric Hansen (SVCW), and Jeromy Miller (Dav). At CDPH, we thank Lauren Baehner, Brooke Bregman, David Rocha, Tony Fristachi and Elana Silver for their help in preparing case data. This study was performed on the ancestral and unceded lands of the Muwekma Ohlone people. We pay our respects to them and their Elders, past and present, and are grateful for the opportunity to live and work here.

